# Impaired semantic control in the logopenic variant of primary progressive aphasia

**DOI:** 10.1101/2024.10.03.24314835

**Authors:** Shalom K. Henderson, Siddharth Ramanan, Matthew A. Rouse, Thomas E. Cope, Ajay D. Halai, Karalyn E. Patterson, James B. Rowe, Matthew A. Lambon Ralph

## Abstract

We investigated semantic cognition in the logopenic variant of primary progressive aphasia (lvPPA), including (i) the status of verbal and non-verbal semantic performance; and (ii) whether the semantic deficit reflects impaired semantic control. Our *a priori* hypothesis that individuals with lvPPA would exhibit semantic control impairments was motivated by the anatomical overlap between the temporoparietal atrophy typically associated with lvPPA and lesions associated with post-stroke semantic aphasia (SA) and Wernicke’s aphasia (WA), which cause heteromodal semantic control impairments. We addressed the presence, type (semantic representation and semantic control; verbal and non-verbal), and progression of semantic deficits in lvPPA. Since most people with lvPPA have Alzheimer’s disease (AD) pathology and are part of a broader multidimensional phenotype space encompassing AD subtypes, we compared semantic performance in lvPPA and typical amnestic AD (tAD). Given the differences in lesion and atrophy patterns in SA and WA *versus* semantic dementia/semantic-variant PPA patients, our second aim was to examine atrophy patterns in people with lvPPA and tAD compared to age-matched controls. Twenty-seven patients participated in the study. People were grouped into those meeting consensus criteria for lvPPA (N = 10) and others who may have previously satisfied definitions of lvPPA but had progressed with multi-domain cognitive impairments (herein referred to as “lvPPA+”; N = 8). People with tAD (N = 9) were relatively preserved across verbal and non-verbal semantic assessments. LvPPA patients were impaired on both verbal and non-verbal semantic tasks and their impairments showed the hallmark characteristics of a semantic control deficit. LvPPA and lvPPA+ patients showed effects of varying semantic control demands, positive cueing effects, and correlated performance between semantic and executive tasks. Whole-brain voxel-based morphometry, comparing each of the patient groups to age-matched controls, revealed significantly reduced grey and white matter in the bilateral hippocampi and lateral temporal regions in tAD patients. The lvPPA group exhibited an asymmetric pattern of reduced grey and white matter intensity in the language-dominant left hemisphere, including a significant portion of the lateral and medial temporal lobe. LvPPA+ patients demonstrated reduced grey and white matter in the left temporal lobe extending subcortically, anteriorly and posteriorly, as well as right temporal involvement. Our findings could aid diagnostic subtyping of PPA by adopting semantic control features and offer improved clinical characterisation of lvPPA in the trajectory of semantic decline.

## Introduction

People’s conceptual knowledge about the world (i.e., semantic representation) and their ability to use and manipulate this information flexibly for a particular context or task (i.e., semantic control) are essential to their overall functional and cognitive status.^1^ Damage to either representation or control systems can be debilitating as found in some types of primary progressive aphasia (PPA) and post-stroke aphasia. Impaired semantic representation (i.e., degraded conceptual knowledge) is a hallmark of the semantic variant of primary progressive aphasia (svPPA)/semantic dementia (SD), due to bilateral atrophy centred on the anterior temporal lobes.^2,3^ Prior studies have contrasted this prominent degradation of semantic representation in svPPA/SD to post-stroke semantic aphasia (SA), where damage to temporoparietal and/or prefrontal cortex impairs semantic control.^4–8^ Despite different lesion profiles (primarily in the posterior superior temporal and supramarginal gyri, extending to posterior middle temporal gyrus), Wernicke’s aphasia (WA) cases have also shown classic features of semantic control impairment including absence of frequency effects and inconsistent performance on various verbal and non-verbal semantic tests when the same items were probed repeatedly.^9,10^

In the current study, we investigated whether semantic cognition is impaired in the logopenic variant of PPA (lvPPA). Although the absence of semantic representation deficits (as observed in SD) constitutes ancillary diagnostic criterion for lvPPA (i.e., spared single-word comprehension and object knowledge),^11^ reports about the type and progression of semantic deficits are mixed^12–16^ (see below) and thus require systematic evaluation across lvPPA at different levels of severity. In addition, the anatomical overlap between the temporoparietal atrophy typically associated with lvPPA and the posterior lesions of the SA and WA patients motivated our *a priori* hypothesis that individuals with lvPPA would exhibit semantic control impairments. It remains unclear if people with lvPPA have poor control of semantic processing across verbal and non-verbal modalities.

We therefore asked three questions: (i) are individuals diagnosed with lvPPA semantically impaired, and if so: (ii) what type of semantic impairment do they have (i.e., semantic representation versus control; in verbal and/or non-verbal domains); and (iii) when does the deficit appear? If individuals with lvPPA exhibit a semantic deficit, this could aid diagnostic subtyping of PPA by using additional features, such as semantic representation versus control-impaired phenotypes, and improve clinical monitoring of lvPPA patients through a more nuanced documentation of initial to later emerging symptoms in the trajectory of decline.

It is typically reported that semantic representation is intact in lvPPA early in the disease course,^12–14^ but this may be only partial preservation and warrants further investigation. Not only is there a paucity of studies investigating general semantic impairment in lvPPA, but also semantic examination is typically modelled on the studies of svPPA/SD (i.e., single item comprehension and lexical retrieval deficits due to degraded conceptual knowledge). Results regarding the type (i.e., semantic representation versus semantic control) and time course (i.e., initial versus later emerging) of semantic deficits in lvPPA are contradictory in the literature. Some studies report that lexical retrieval deficits in lvPPA (one of the two hallmark features) are underpinned by semantic impairment leading to production problems.^15,17–20^ Given the wide usage of single word comprehension tasks to detect degraded conceptual knowledge in svPPA/SD, Leyton *et al*. reported that naming deficits in some individuals with lvPPA are due to impaired semantic processing as they exhibited poor performance on tests of picture naming and single word comprehension.^21^ Similarly, Galton *et al*. found that among individuals with Alzheimer’s disease (AD) who presented with a predominant language profile (i.e., predating the lvPPA classification), over 80% demonstrated semantic deficits during in depth neuropsychological testing.^22^ Intragroup variability poses another challenge as the degree of semantic deficit can vary from one individual to another. For example, Migliaccio *et al*. noted that mean scores on a single word comprehension test in the lvPPA group were below the published normative cut-off because two patients had significantly greater word comprehension deficits than the rest.^23^

Features of semantic control problems, of the form observed in SA and WA, may be gleaned from recent studies showing positive effects of phonemic cueing and phonological facilitation of related but not unrelated words in individuals with lvPPA.^7,24^ Semantic interference effects have also been reported in lvPPA, where reaction times were found to be slower for semantically related, but not for unrelated, words.^25^ An alternative hypothesis is that individuals with lvPPA exhibit both representation and control deficits. This idea is in line with Corbett e*t al.* who showed that the nature of semantic impairment in AD was modulated by disease severity; mild AD patients presented with control deficits and those with severe AD presented with additional representation deficits.^26^ Given that the majority of lvPPA patients have AD pathology^27–29^ and can be considered to be a part of a broader graded multidimensional phenotype space between AD subtypes,^30–33^ comparing individuals with lvPPA to those with typical AD (i.e., with a predominant amnestic presentation) facilitates a direct comparison of semantic deficits across typical and atypical presentations of AD.

Classic atrophy patterns in lvPPA are centred on the left temporoparietal junction, specifically the superior and middle temporal gyri and the inferior parietal lobule,^11,34^ and thus overlap with regions of the semantic control network. The lateral posterior temporal cortex, particularly the posterior middle and inferior temporal gyri, and the inferior frontal gyrus are key regions of the semantic control network in both patients and healthy participants.^35,36^ Previous studies have highlighted the heterogeneous nature of atrophy in lvPPA. The distribution and extent of atrophy in those with lvPPA can vary substantially and may include the superior parietal, anterior temporal, and inferior frontal regions.^31,32,37,38^ Additionally, only a few studies have provided information on the longitudinal pattern of semantic decline in lvPPA. In a voxel-based morphometry analysis of lvPPA patients, Rohrer *et al.* found that at initial assessment the most significant areas of atrophy encompassed the left superior and middle temporal gyri, inferior parietal, and medial temporal lobe.^39^ Longitudinal imaging and neuropsychological analyses revealed increasing involvement of more anterior and medial temporal lobe regions, particularly the superior temporal gyrus, and worsening performance on naming, as well as single word and sentence comprehension. The occurrence of semantic deficits, however, may not be only associated with atrophy encroaching on the anterior temporal lobe. Funayama *et al*. reported that lvPPA patients who progressed to have greater middle and posterior temporal involvement of the inferior temporal gyrus exhibited svPPA-like semantic memory deficits (see the authors’ Case 1).^16^ Schaeverbeke *et al*. postulated that semantic deficits may be (i) due to the extension of damage into the posterior third of the superior temporal sulcus and the middle temporal gyrus and (ii) related to disturbances in top-down semantic control: their sample of mixed variant of PPA who made the most errors on trials assessing the non-dominant meaning of homonyms during a single word comprehension task.^40^ Thus, examination of the spatial extent and the distribution of atrophy in lvPPA patients across different stages of severity could help to elucidate whether semantic impairment might be modulated by disease severity.

This study sought to address the uncertainty about the presence, type and progression of verbal and non-verbal semantic deficits in lvPPA. We assessed performance on various verbal and non-verbal semantic tests and examined atrophy patterns in patients with lvPPA and more typical, amnestic AD, compared to age-matched healthy controls. We tested the hypotheses that if individuals with lvPPA exhibit a semantic deficit, they will show (a) varying performance according to semantic control demands across verbal and non-verbal task modalities, (b) no or very limited influence of frequency and familiarity, (c) positive effects of phonemic cueing, and (d) correlated performance for semantic control and executive tasks.

## Materials and methods

### Participants

People with clinical diagnoses of lvPPA or Alzheimer’s disease (N = 27) were recruited from specialist clinics for memory disorders within the Cambridge University Hospitals. Twelve healthy controls were recruited from the MRC Cognition and Brain Sciences Unit volunteer panel. Patient participants completed a comprehensive clinical evaluation including a multidisciplinary assessment, full clinical history with patient and next of kin, structured neurological, neuroimaging, and cognitive examinations. All participants self-reported as “White” and reported English as their first language. At the time of study participation, nine patients had typical, amnestic presentation of AD (tAD),^41^ ten patients met strict criteria for lvPPA,^11,42^ and eight patients were classified as “lvPPA+” as they previously satisfied definitions of lvPPA but, at the time of this study, they exhibited multi-domain cognitive impairments. The lvPPA+ patients in our sample facilitated an examination of the multidimensional cognitive impairments across lvPPA disease severity. All participants gave written informed consent in accordance with the Declaration of Helsinki, with patients being supported by family when necessary.

Out of the semantic assessments listed below, only one lvPPA+ patient did not complete the Camel and Cactus Test and the synonym judgement task due to difficulty understanding task instructions, and the alternative object use task was not administered to three patients either due to time constraints or other personal factors. All other participants completed all of the semantic assessments. For a few participants, some of the additional executive tasks and the cued verbal fluency tasks were discontinued due to task difficulty, time constraints, and/or other personal factors, and details about the missing executive data are summarised in the statistical analysis section.

### Assessments

#### General neuropsychology

General neuropsychological assessments were administered to participants, including the Addenbrooke’s Cognitive Examination – Revised (ACE-R),^43^ forwards and backwards digit span from the Wechsler Memory Scale – Revised,^44,45^ Trail Making Test,^46^ the Raven’s Coloured Progressive Matrices test of non-verbal reasoning,^47^ Hayling and Brixton tests.^48^ Patients’ next of kin completed the revised Cambridge Behavioural Inventory questionnaire.^49^

#### Semantic cognition assessment

Consistent with previous investigations of semantic control,^4,7,8^ semantic cognition was assessed using (1) 30-item Boston Naming Test (BNT), which was used not only to assess total pictures named spontaneously, but also additional names produced following phonemic cues^50^; (2) 64-item Cambridge Semantic Battery (CSB), which probed the same items in three subtests consisting of picture naming, spoken word-picture matching, and the picture version of the Camel and Cactus Test (CCT)^51^; (3) 48-item synonym judgement task, where the participant was asked to match a probe word to a synonym target presented with two unrelated distractors^52,53^; (4) 37-item alternative object use task, which assessed the canonical and alternative uses of everyday objects (e.g., a ‘fly swat’ would be a canonical object to kill a fly whereas a rolled-up newspaper would constitute an alternative object) with either semantically related or unrelated distractors^54^; and (5) the 16-item spoken sentence-to-picture matching subtest of the Comprehensive Aphasia Test (CAT), which assessed each participant’s language comprehension of sentences.^55^

Participants completed a standard test of verbal fluency, where they produced as many words as possible within one minute for the category of ‘animals’ and the letter ‘P’. In addition, we administered a cued semantic fluency task where the categories of ‘animals’ and ‘supermarket’ were each divided into four 15-second blocks with the provision of subcategory cues (e.g., for animals: animals that people keep in their homes as pets, animals that are found on a farm, animals that live in the jungle, and animals that live in water; for supermarket: fruits and vegetables, meat and seafood, things people drink, and household cleaning products).^56–58^ We designed a letter fluency task similar to Song *et al*. and asked the participants to name as many words as possible that began with ‘fa’, ‘fo’, ‘fl’, and ‘fr’ in 15-second blocks.^59^ The instructions were as follows: “Please name as many words as possible that start with the letters (e.g., F, A). I will stop you after 15 seconds”. Along with the verbal instruction, a written prompt with the two letter combinations was also provided as support. The examiner (SKH) did not sound out the phonemes (e.g., fæ) as this would further constrain the task to a smaller set of word choices available for the F words followed by a vowel (e.g., /fæ/ for words like “fat” and “fan”, and /feI/ for words like “fail” and “faint”).

### Statistical analysis

To assess performance across semantic tasks across all groups (i.e., controls, tAD, lvPPA, lvPPA+), we conducted a Bayesian ANOVA to test for group differences in each semantic task followed by a *post hoc* test to indicate the adjusted posterior model odds and Bayes factors (BFs). Unless otherwise specified, controls were excluded from the analyses as they exhibited ceiling effects.

Using the alternative object use task, performance on semantic control was further examined with 2 canonicity (i.e., canonical versus alternative condition) x 2 distractor (i.e., semantically related versus unrelated) Bayesian repeated measures ANOVA with group as a between subjects factor.

We assessed the effect of cueing in two ways. To measure how patient participants were aided by phonemic cues on the BNT, we compared the uncued (i.e., total items named spontaneously) and cued (i.e., total items named spontaneously plus with phonemic cues) performance by conducting a 2 condition (i.e., uncued, cued) x 3 groups (i.e., tAD, lvPPA, lvPPA+) Bayesian ANOVA. Second, we compared the uncued and cued category (i.e., performance on cued ‘animals’ and ‘supermarket’ averaged together) and letter fluency with a 2 condition (i.e., uncued, cued) x 2 fluency type (i.e., letter, category) x 4 groups (including controls) Bayesian ANOVA. Controls were included in this analysis because (1) there is no maximum score for the test of verbal fluency, and (2) literature is sparse on whether controls might benefit from cueing on fluency performance.

Previous studies of SA have shown that performance on more demanding semantic tasks is highly correlated with performance on executive function tests such as the Raven’s Coloured Progressive Matrices and digit span backward.^4,60^ To this end, we first imputed the missing data for executive tasks (14% overall) using estim_ncpPCA in R (https://www.rdocumentation.org/packages/missMDA/versions/1.19/topics/estim_ncpPCA), a widely used method to impute data with cross-validated principal component analysis (PCA) per previous protocols.^61,62^ Next, we performed a constrained, varimax-rotated PCA with the executively-tapping tasks from the detailed neuropsychology battery: namely the Raven’s Coloured Progressive Matrices, digit span backward, Trail making test B, and Brixton, to derive a single principal component score per participant that is representative of his or her overall executive functioning performance. We examined the association between the ‘executive’ principal component score and accuracy on the three semantic tasks that have been previously shown to tax semantic control demands: CCT, alternative object use, and synonym judgement task. Given the normality of data, we used Pearson’s *rho* correlation and report the associated Bayes factors.

### Neuroimaging acquisition and analysis

Participants (12 healthy controls, 27 patients) completed a T1-weighted 3T structural MRI scan on a Siemens PRISMA at the University of Cambridge (GRAPPA acceleration factor = 2). Thirty-five participants were scanned at the MRC Cognition and Brain Sciences Unit with the following parameters: sagittal image acquisition, no. slices = 208, TR = 2000ms, TE = 2.85mg, flip angle = 8°, FOV = 228.8 x 281.6 x 281.6mm^3^, resolution matrix = 208 x 256 x 256, voxel size = 1.1mm^3^. Four participants were scanned at the Wolfson Brain Imaging Centre with the following parameters: sagittal image acquisition, no. slices = 208, TR = 2000ms, TE = 2.93, flip angle = 8°, FOV = 228.8 x 281.6 x 281.6mm^3^, resolution matrix = 228.8 x 256 x 256, voxel size = 1.1mm^3^.

The T1-weighted MPRAGE images were preprocessed using the processing stream of the Computational Anatomy Toolbox 12 (CAT12) (https://www.neuro.uni-jena.de/cat/) in the Statistical Parametric Mapping software (SPM12: Wellcome Trust Centre for Neuroimaging, https://www.fil.ion.ucl.ac.uk/spm/software/spm12/). Our pre-processing pipeline used: (i) denoising, resampling, bias-correction, affine registration, and brain segmentation into three tissue probability maps (grey matter, white matter, cerebrospinal fluid); (ii) normalisation and registration to the Montreal Neurological Institute (MNI) template, and (iii) smoothing using 8mm full-width-half-maximum Gaussian kernel. Segmented, normalised, modulated, and smoothed grey and white matter images were used for voxel-based morphometry (VBM) analysis.

Consistent with previous studies,^31,63^ we used grey and white matter VBM to account for co-occurring grey and white matter changes that are typical of lvPPA.^64^ We included 27 additional age-matched control participant scans from the Cambridge Centre for Frontotemporal Dementia database. Voxel-wise differences of grey and white matter intensity between patients *versus* control groups were assessed using independent *t*-tests, with age and total intracranial volume included as nuisance variables. Clusters were extracted, corrected for Family-Wise Error (FWE) at *P* < 0.05, as well as for False Discovery Rate (FDR) at *P* < 0.001, with a cluster threshold of 100 contiguous voxels. We employed the additional lenient FDR threshold because the FWE correction method may be too stringent given the amount of atrophy in patients with neurodegenerative diseases.

## Results

### Demographics

Demographic details are shown in Table 1. Bayesian ANOVA revealed no evidence for differences in all groups for age and handedness (*P* > 0.05, BF < 0.33) and there was no evidence for a difference in gender (*P* > 0.05, BF = 0.54). The results of a Bayesian ANOVA showed anecdotal evidence for a difference in self-reported symptom duration for patients (*P* = 0.05, BF = 1.66), which was driven by patients with tAD having longer symptom duration than those with lvPPA (*P* = 0.04, BF = 3.36). The anecdotal evidence for a difference in education (*P* = 0.05, BF = 1.91) was driven by controls having somewhat higher levels of education than patients, but the results of pairwise multiple comparisons did not reveal any differences between controls and each of the patient groups, and patient groups also did not differ from one another (*P* > 0.05, BF < 2.00).

**Table 1.** Demographics and clinical features of the study cohort.

|  | **Control** | **tAD** | **lvPPA** | **lvPPA+** | ***P*\*** | ***BF*** |
| --- | --- | --- | --- | --- | --- | --- |
| N | 12 | 9 | 10 | 8 | - | - |
| Age (years) | 71.08<br>(8.89) | 74.56<br>(6.48) | 72.870<br>(8.98) | 68.63<br>(5.64) | ns | 0.31 |
| Gender (male/female) | 5/7 | 6/3 | 5/5 | 7/1 | ns | 0.54 |
| Handedness (right/left) | 10/2 | 8/1 | 9/1 | 8/0 | ns | 0.20 |
| Symptom duration (years) | - | 5.11<br>(2.32) | 3.00<br>(1.05) | 4.00<br>(1.85) | 0.05 | 1.66 |
| Mean age at leaving full-time education (years) | 21.5<br>(6.47) | 17.44<br>(2.46) | 16.70<br>(3.06) | 17.38<br>(2.39) | 0.05 | 1.91 |
| Mean Cambridge Behavioural Inventory – Revised (maximum score 180) | 4.33<br>(7.50) | 57.00<br>(41.43) | 31.30<br>(19.20) | 48.38<br>(34.38) | < 0.001 | 70.11 |
| Mean ACE-R (maximum score 100) | 94.33<br>(5.58) | 77.44<br>(9.03) | 60.90<br>(9.61) | 37.71<br>(17.40) | < 0.001 | >100 |
| Mean ACE-R language subdomain (maximum score 26) | 25.83<br>(0.58) | 25.00<br>(1.12) | 16.60<br>(3.98) | 12.71<br>(6.32) | < 0.001 | >100 |
| Mean ACE-R memory subdomain (maximum score 26) | 22.58<br>(3.78) | 15.38<br>(4.69) | 12.90<br>(4.51) | 4.00<br>(3.95) | < 0.001 | >100 |

Note: Mean and standard deviations are displayed. \**P*-value for *F*-test of group-difference by ANOVA. ACE-R, Addenbrooke’s Cognitive Examination Revised; BF, Bayes factor; lvPPA, logopenic variant primary progressive aphasia; ns, not significant; tAD, typical Alzheimer’s disease.

Pairwise multiple comparisons confirmed very strong to extreme evidence that controls had significantly lower scores on the Cambridge Behavioural Inventory compared to all three patient groups (BF > 50), but patient groups did not differ from one another. As expected, evidence for a difference in ACE-R total scores between controls and all three patient groups was extreme (BF > 100). Across the patient groups, evidence was extreme between tAD and lvPPA+ (*P* < 0.001, BF > 100), and strong between tAD and lvPPA (*P* = 0.007, BF = 24.72) and lvPPA and lvPPA+ (*P* < 0.001, BF = 12.69). For ACE-R language sub-scores, there was extreme evidence for a difference between controls and tAD versus lvPPA and lvPPA+ (*P* < 0.001, BF > 100). For the subdomain of memory, evidence for a difference ranged from moderate for controls versus tADs (*P* = 0.004, BF = 12.69) to extreme for controls versus lvPPA and lvPPA+ (*P* < 0.001, BF > 100). Evidence for a difference in ACE-R memory sub-scores was also very strong between tAD and lvPPA+ (*P* < 0.001, BF = 80.15).

### Behavioural results

#### Performance across semantic tasks

Bayesian ANOVAs comparing group performance across each semantic task showed evidence of a group effect. Group performance patterns on each semantic task are visually summarised in Figure 1 and Supplementary Table 1 shows the adjusted posterior odds. There was extreme evidence in favour of a group effect for the BNT (F(3,35) = 68.18, *P* < 0.001, BF > 100) with *post hoc* tests showing evidence that performance differed between controls and all patient groups (BF > 100). The evidence for the group difference was extreme for tAD versus lvPPA+ and lvPPA (*P* < 0.001, BF > 100), and moderate between lvPPA and lvPPA+ (*P* = 0.001, BF = 5.19).

**Figure 1.**
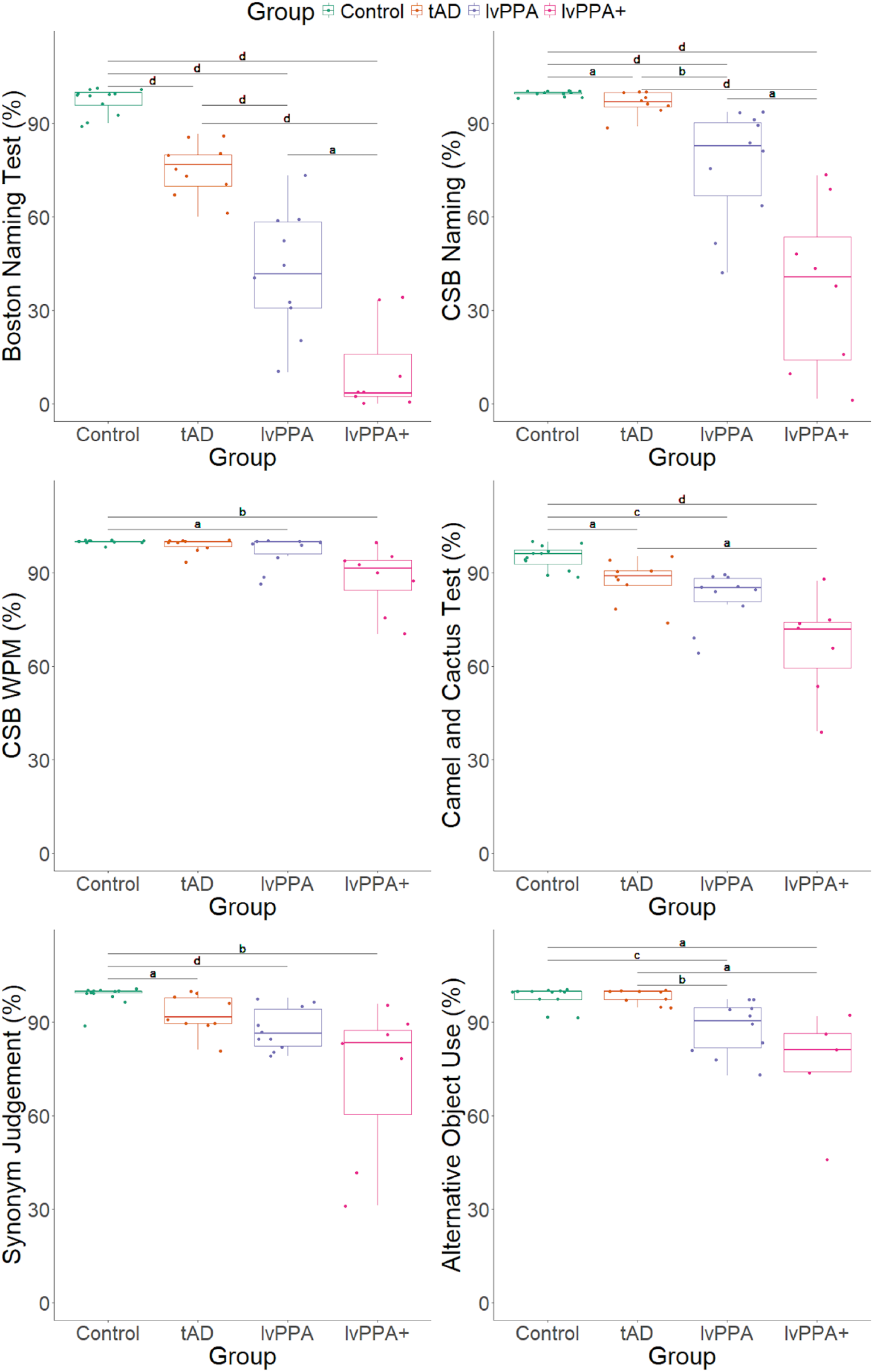
Boxplots showing group performance across semantic tasks. Boston Naming Test (*top left*), Cambridge Semantic Battery (CSB) naming subtest *(top right*), CSB word-picture matching (WPM) subtest (*middle left*), Camel and Cactus Test (*middle right*), synonym judgement task (*bottom left*), and alternative object use task (*bottom right*). Bayesian ANOVAs tested for group differences (Control N = 12; tAD N = 9; lvPPA N = 10; lvPPA+ N = 8) in each semantic task and results from *post hoc* group comparisons are shown as letters indicating level of evidence: a = moderate (3 < Bayes Factor (BF) < 10); b = strong (10 ≤ BF < 30); c = very strong (30 ≤ BF < 100); d = extreme (BF > 100). Each dot represents individual performance scores across the six semantic tasks. Note: A single lvPPA+ patient did not complete the Camel and Cactus Test and the synonym judgement task, and three lvPPA+ patients did not complete alternative object use task. lvPPA, logopenic variant primary progressive aphasia; tAD, typical Alzheimer’s disease.

For CSB naming, there was extreme evidence in favour of a group effect (F(3,35) = 27.45, *P* < 0.001, BF > 100) with *post hoc* tests showing that performance differed between controls and all patient groups, with evidence also being extreme for controls versus lvPPA and lvPPA+ (*P* < 0.01, BF > 100). There was moderate evidence for a difference between controls and tAD (*P* = 0.98, BF = 4.26). Across the patient groups, evidence for a group difference was extreme between tAD and lvPPA+ (*P* < 0.001, BF > 100), strong between tAD and lvPPA (*P* = 0.03, BF = 12.70), and moderate between lvPPA and lvPPA+ (*P* < 0.001, BF = 9.37).

There was strong evidence in favour of a group effect for CSB WPM (F(3,35) = 6.06, *P* = 0.002, BF = 18.63), particularly between controls and lvPPA+ (*P* = 0.002, BF = 11.20). Evidence for a difference was anecdotal between tAD and lvPPA+ (*P* = 0.009, BF = 2.91) and moderate between controls and lvPPA (*P* = 0.40, BF = 3.08).

For CCT, there was extreme evidence in favour of a group effect (F(3,34) = 12.76, *P* < 0.001, BF > 100) with *post hoc* tests showing that performance differed between controls and all of the patient groups, with evidence being extreme for controls versus lvPPA+ (*P* < 0.001, BF > 100), very strong versus lvPPA (*P* = 0.02, BF = 92.50), and moderate versus tAD (*P* = 0.25, BF = 9.96). Across patient groups, the evidence for the difference was moderate between tAD and lvPPA+ (*P* = 0.002, BF = 6.71) and anecdotal for lvPPA and lvPPA+ (*P* = 0.04, BF = 1.64).

For the synonym judgement task, there was very strong evidence in favour of a group effect (F(3,34) = 7.71, *P* < 0.001, BF= 65.47) with strong evidence that performance differed between controls and lvPPA+ (*P* < 0.001, BF = 20.24). *Post hoc* tests revealed evidence that was extreme for controls versus lvPPA (*P* = 0.24, BF > 100) and moderate versus tAD (*P* = 0.68, BF = 4.84). In the patient groups, evidence for difference was moderate between tAD and lvPPA+ (*P* = 0.009, BF = 2.40) and anecdotal between lvPPA and lvPPA+ (*P* = 0.05, BF = 1.27).

Finally, evidence in favour of a group effect was very strong for the alternative object use task (F(3,31) = 7.26, *P* < 0.001, BF = 36.32). Evidence was very strong between controls and lvPPA (*P* = 0.03, BF = 56.83), strong between tAD and lvPPA (*P* = 0.05, BF = 21.39), moderate between controls and lvPPA+ (*P* = 0.003, BF = 6.07), and moderate between tAD and lvPPA+ (*P* = 0.005, BF = 3.17).

#### Performance on semantic control-demanding alternative object use task

As shown in Figure 1, controls were mostly at ceiling for the alternative object use task and were excluded in this analysis. Bayesian ANOVA revealed extreme evidence for an effect of canonicity (F(1,21) = 29.27, *P* < 0.001, BF > 100), strong evidence for the effects of distractor (F(1,21) = 10.04, *P* = 0.005, BF = 20.26) and group (F(2,21) = 6.48, *P* = 0.006, BF = 12.07), as well as moderate to strong evidence for canonicity-by-distractor (F(1,21) = 6.46, *P* = 0.02, BF = 16.67) and canonicity-by-group (F(1,21) = 4.42, *P* = 0.03, BF = 4.04) interactions. Across the whole group, *post hoc* tests revealed strong evidence that performance differed between canonical and alternative types (BF > 100) as shown in Figure 2; evidence was moderate between semantically related versus unrelated distractor types (BF = 6.85). Group comparisons showed that performance differed between patient groups, with evidence being extreme between tAD and lvPPA+ (BF > 100) and strong between tAD and lvPPA (BF = 20.33).

**Figure 2.**
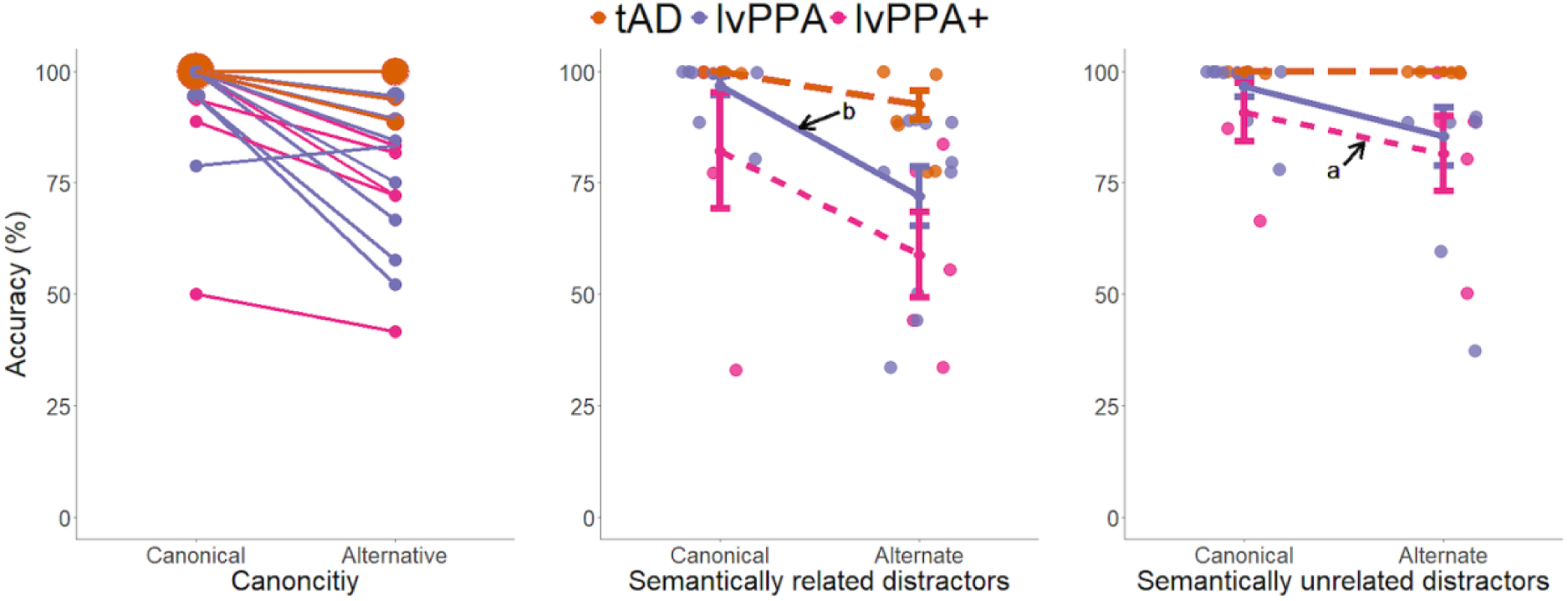
Performance on the alternative object use task. Individual patterns for ‘canonical’ and ‘alternative’ conditions (*left*), where the larger dots denote more participants with the same score, overall group patterns for ‘canonical’ and ‘alternative’ conditions with semantically related distractors (*middle*) and with unrelated distractors (*right*) with standard error of the mean. Each dot represents individual performance scores (tAD N = 9; lvPPA N = 10; lvPPA+ N = 5). Bayesian ANOVA revealed extreme evidence for an effect of canonicity (F(1,21) = 29.27, *P* < 0.001, BF > 100), strong evidence for the effects of distractor (F(1,21) = 10.04, *P* = 0.005, BF = 20.26) and group (F(2,21) = 6.48, *P* = 0.006, BF = 12.07), as well as moderate to strong evidence for canonicity-by-distractor (F(1,21) = 6.46, *P* = 0.02, BF = 16.67) and canonicity-by-group (F(1,21) = 4.42, *P* = 0.03, BF = 4.04) interactions. Results from *post hoc* analyses comparing performance in canonical versus alternative and semantically related versus unrelated distractor conditions are shown as letters indicating level of evidence: a = moderate (3 < Bayes Factor (BF) < 10); b = strong (10 ≤ BF < 30); c = very strong (30 ≤ BF < 100); d = extreme (BF > 100). lvPPA, logopenic variant primary progressive aphasia; tAD, typical Alzheimer’s disease.

We conducted *post hoc* Bayesian paired sample *t*-tests within each group to compare performance in canonical (C) versus alternative (A) and semantically related (S) versus unrelated (U) distractor conditions, as well as across all condition comparisons (i.e., CU-AU, CS-AS, CU-CS, AU-AS). Patients in the tAD group were mostly at ceiling resulting in zero variance for the *t*-tests. In the lvPPA group, evidence that performance was lower for (i) the alternative relative to canonical condition overall was moderate (*t* = 3.71, *P* = 0.005, BF = 11.15) and (ii) AS relative to CS condition was moderate (*t* = 3.72, *P* = 0.005, BF = 11.29). In the lvPPA+ group, there was moderate evidence that performance was lower for (i) the alternative relative to canonical condition overall (*t* = 4.99, *P* = 0.008, BF = 8.66) and (ii) AU relative to CU condition (*t* = 3.38, *P* = 0.03, BF = 3.38).

#### Effect of familiarity/frequency

A 4 task x 2 familiarity Bayesian repeated measures ANOVA with group as a between subjects factor (excluding controls who were at ceiling) revealed strong evidence in favour of an effect of familiarity/frequency (F(1,69) = 21.83, *P* < 0.001, BF = 15.73), but no evidence for the interactions between familiarity/frequency and task, group, and task and group (*P* > 0.05, BF < 1). *Post hoc* comparisons showed evidence that overall performance across all groups differed between high versus low familiarity/frequency items (BF > 100). Figure 3 shows the results for the three patient groups and includes the figures and results from Jefferies and Lambon Ralph (2006) and Thompson *et al.* (2018) where they have conducted the same analyses with SD versus SA patients.^4,8^

**Figure 3.**
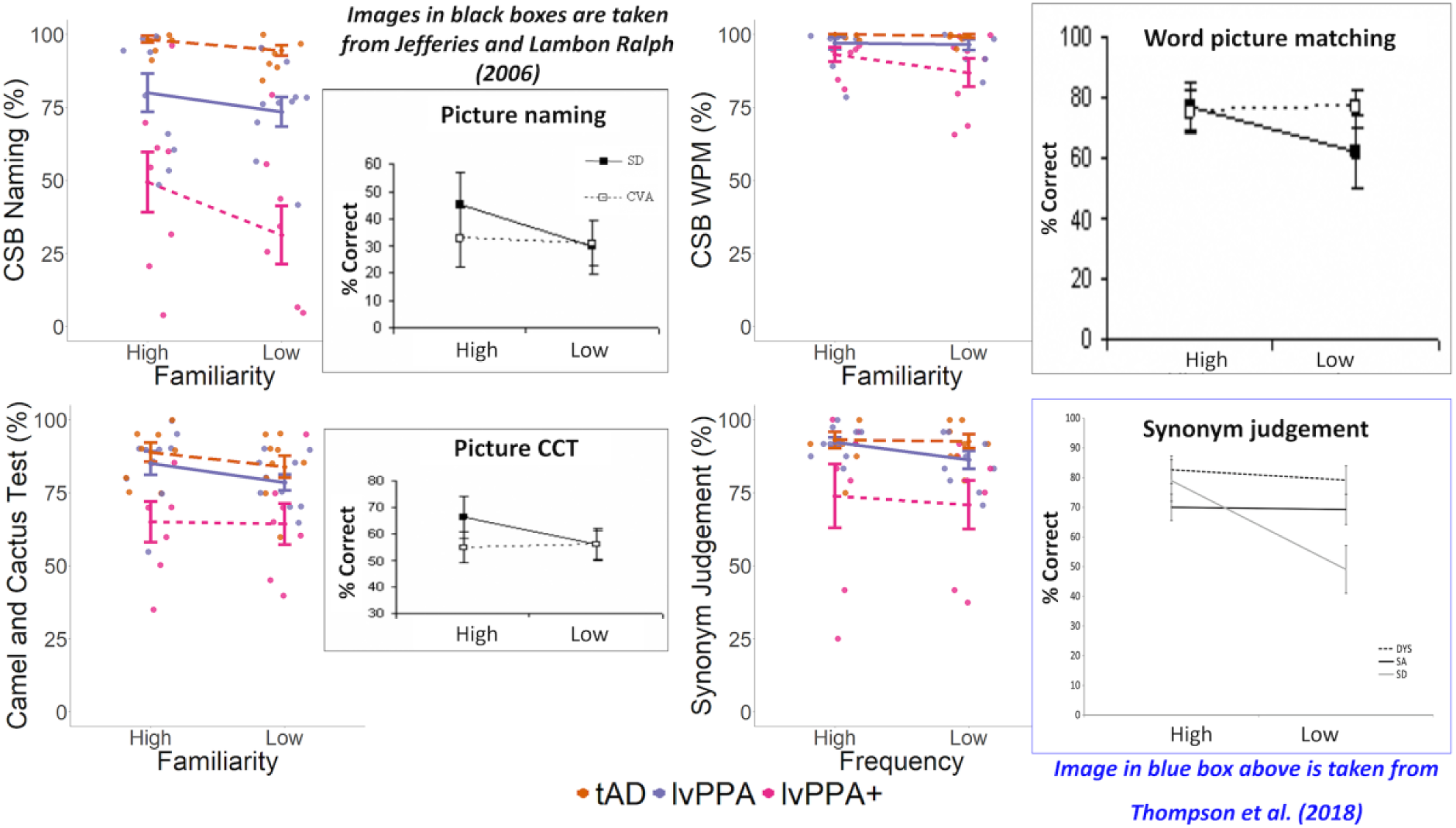
Effect of familiarity/frequency on the Cambridge Semantic Battery (CSB) subtests and the synonym judgement task. Overall group patterns (tAD N = 9; lvPPA N = 10; lvPPA+ N = 8) for high versus low familiarity/frequency items on the CSB naming (*top left*), CSB WPM (*top right*), Camel and Cactus Test (*bottom left*), and the synonym judgement task (*bottom right*) with standard error of the mean. The results of a Bayesian repeated measures ANOVA revealed strong evidence in favour of an effect of familiarity/frequency (F(1,69) = 21.83, *P* < 0.001, BF = 15.73), but no evidence for the interactions between familiarity/frequency and task, group, and task and group (*P* > 0.05, BF < 1). Each dot represents individual performance scores. The figures in black boxes were taken from Jefferies and Lambon Ralph (2006) including subjects with Semantic Dementia (SD) and semantic aphasia (SA) following cerebrovascular accident (CVA) and the figure in the blue box was taken from Thompson *et al.* (2018) including subjects with SD, SA, and dysexecutive syndrome (DYS) on the same tests as the present study for visual comparison. Note: A single lvPPA+ patient did not complete the Camel and Cactus Test and the synonym judgement task. lvPPA, logopenic variant primary progressive aphasia; tAD, typical Alzheimer’s disease; WPM, word-picture matching.

#### Effect of cueing

Excluding controls, the results of the Bayesian ANOVA assessing the effect of phonemic cueing on BNT performance revealed extreme evidence in favour of an effect of cueing (F(1,48) = 33.90, *P* < 0.001, BF > 100). There was anecdotal evidence for a cueing-by-group interaction (F(2,48) = 1.36, *P* = 0.27, BF = 2.07). *Post hoc* paired samples *t*-tests revealed extreme evidence for a cueing effect for all patient groups (BF > 100).

The results of the Bayesian ANOVA assessing the effect of cueing on verbal fluency performance revealed moderate evidence for cueing (F(1,102) = 4.67, *P* = 0.03, BF = 4.57) and an extreme effect of fluency type (F(1,102) = 9.80, *P* = 0.002, BF > 100), which was driven by more words produced for cued relative to uncued condition, as well as for category relative to letter fluency, as shown in Figure 4. There was no evidence for an interaction between cueing and fluency type, group, and fluency and group (*P* > 0.05, BF < 2).

**Figure 4.**
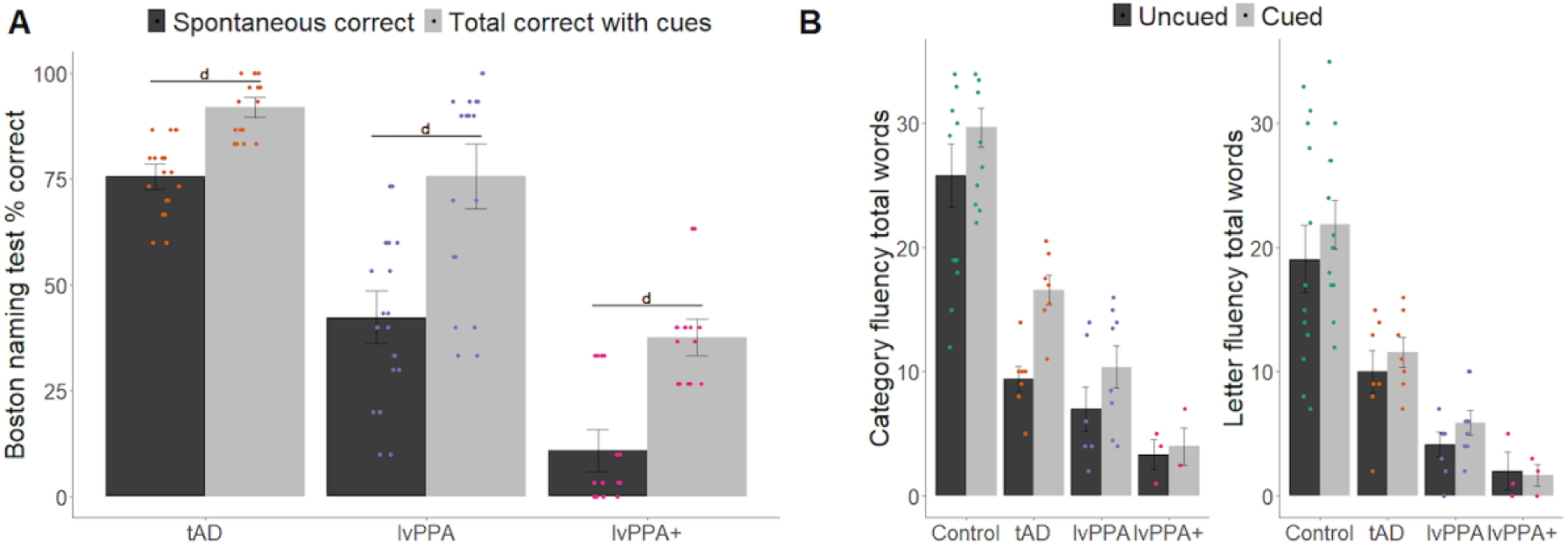
Effect of cueing on (A) picture naming and (B) verbal fluency. Overall group patterns are shown with standard error of the mean. For picture naming (tAD N = 9; lvPPA N = 10; lvPPA+ N = 8), the results of the Bayesian ANOVA revealed extreme evidence in favour of an effect of cueing (F(1,48) = 33.90, *P* < 0.001, BF > 100) and anecdotal evidence for a cueing-by-group interaction (F(2,48) = 1.36, *P* = 0.27, BF = 2.07). *Post hoc* paired samples *t*-tests revealed extreme evidence for a cueing effect for all patient groups (BF > 100). For verbal fluency (Control N = 12; tAD N = 7; lvPPA N = 8; lvPPA+ N = 3), the results of the Bayesian ANOVA revealed moderate evidence for cueing (F(1,102) = 4.67, *P* = 0.03, BF = 4.57) and an extreme effect of fluency type (F(1,102) = 9.80, *P* = 0.002, BF > 100). There was no evidence for an interaction between cueing and fluency type, group, and fluency and group (*P* > 0.05, BF < 2). Results from *post hoc* analyses are shown as letters indicating level of evidence: a = moderate (3 < Bayes Factor (BF) < 10); b = strong (10 ≤ BF < 30); c = very strong (30 ≤ BF < 100); d = extreme (BF > 100), and each dot represents individual performance scores. lvPPA, logopenic variant primary progressive aphasia; tAD, typical Alzheimer’s disease.

#### Correlations between semantic and executive tasks

A single ‘executive’ principal component (PC) score was derived for each participant from a constrained, varimax-rotated principal component analysis which explained 61% of the variance (Kaiser-Meyer-Olkin = 0.61). The loadings of each executive measure are shown in Supplementary Table 2.

As shown in Figure 5, evidence for a correlation between ‘executive’ PC scores and performance on the CCT was extreme in the whole group (*r* = 0.66, *P* < 0.001, BF > 100), very strong in the lvPPA/lvPPA+ combined group (*r* = 0.73, *P* < 0.001, BF = 50.05), and anecdotal in the lvPPA group (*r* = 0.64, *P* = 0.05, BF = 2.16). There was no evidence for a correlation between ‘executive’ PC scores and performance on the alternative object use task in all groups (*P* > 0.05, 0.33 < BF < 2). Evidence for a correlation between ‘executive’ PC scores and performance on the synonym judgement task was strong in the whole group (*r* = 0.59, *P* = 0.002, BF = 29.24) and moderate in the lvPPA/lvPPA+ combined group (*r* = 0.57, *P* = 0.02, BF = 4.20). This correlation was not observed in the other groups (0.33 < BF < 3).

**Figure 5.**
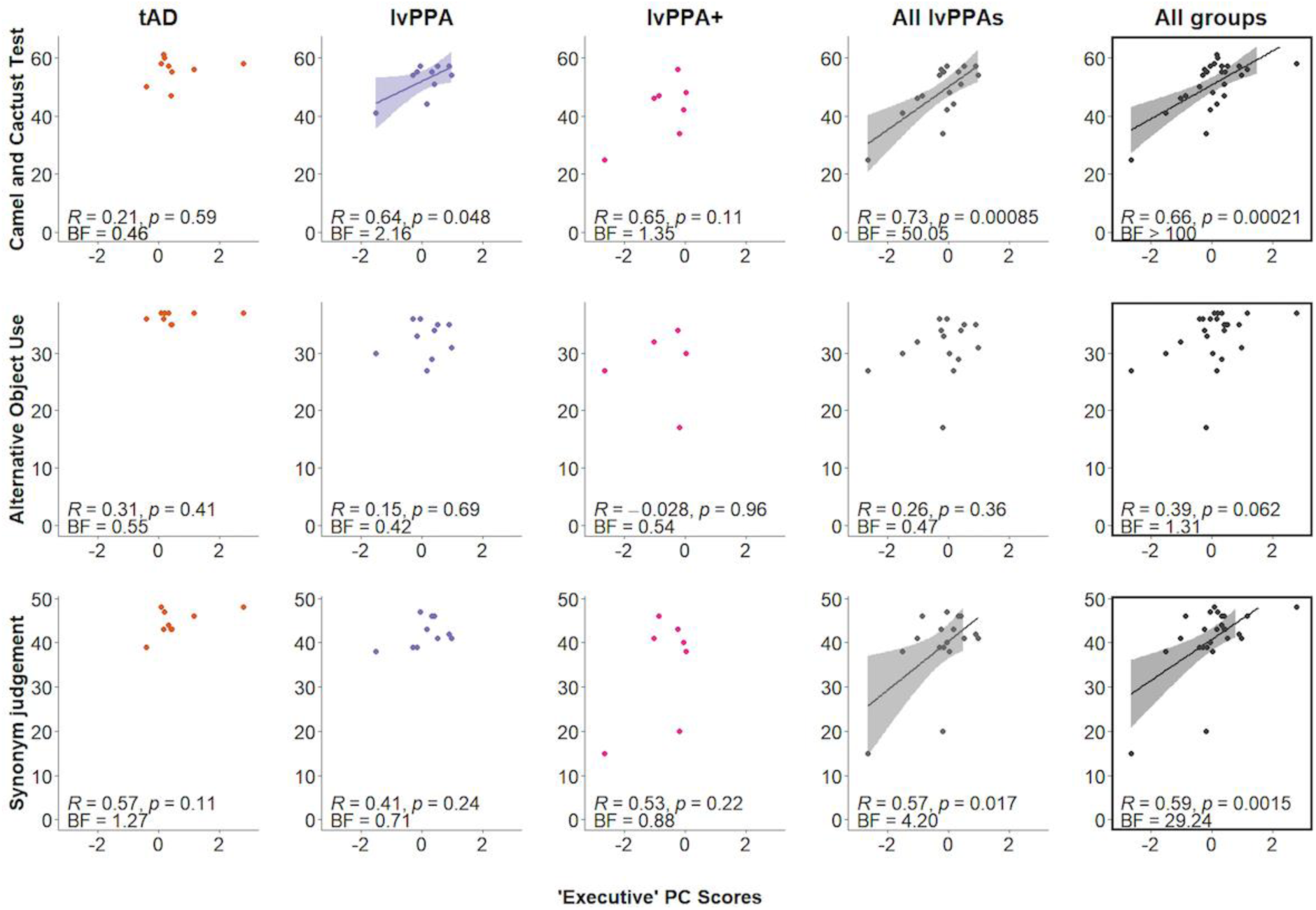
Associations between ‘executive’ principal component scores and three semantic controls tasks across all patient groups. The x-axis represents the ‘executive’ principal component (PC) scores. The y-axis represents the raw scores for each test and the maximum scores are 64 for the Camel and Cactus Test (tAD N = 9; lvPPA N = 10; lvPPA+ N = 7), 37 for the alternative object use task (tAD N = 9; lvPPA N = 10; lvPPA+ N = 5), and 48 for the synonym judgement task (tAD N = 9; lvPPA N = 10; lvPPA+ N = 7). The scatterplots entitled “All lvPPAs” illustrate the combined lvPPA and lvPPA+ group data. Evidence for a correlation between ‘executive’ PC scores and performance on the CCT was extreme in the whole group (*r* = 0.66, *P* < 0.001, BF > 100), very strong in the lvPPA/lvPPA+ combined group (*r* = 0.73, *P* < 0.001, BF = 50.05), and anecdotal in the lvPPA group (*r* = 0.64, *P* = 0.05, BF = 2.16). Evidence for a correlation between ‘executive’ PC scores and performance on the synonym judgement task was strong in the whole group (*r* = 0.59, *P* = 0.002, BF = 29.24) and moderate in the lvPPA/lvPPA+ combined group (*r* = 0.57, *P* = 0.02, BF = 4.20). BF, Bayes factor; lvPPA, logopenic variant primary progressive aphasia.

### Neuroimaging results

Differences in grey and white matter in the patient groups relative to controls are displayed in Figure 6. As shown in Supplementary Table 3, when using *P* < 0.05 FWE-correction, patients had significantly reduced grey and white matter intensity in the left temporal lobe, right middle frontal gyrus, parietal, postcentral and superior temporal gyri relative to controls. The tAD group had significantly reduced grey and white matter intensity in the bilateral hippocampi, medial and lateral temporal, and medial frontal regions, as well as the insula. The lvPPA group showed asymmetric left-lateralised grey and white matter intensity in the left temporal lobe extending subcortically including the hippocampus, fusiform and parahippocampal gyri, and posteriorly into the temporo-parietal and occipital regions, as well as smaller clusters in the right middle, inferior, and fusiform gyri, and the medial frontal gyrus. The lvPPA+ group further demonstrated reduced grey and white matter intensity in the left temporal lobe extending subcortically, anteriorly, and posteriorly into the parietal lobule. Relative to the lvPPA group map, the lvPPA+ group also showed involvement of right hemisphere regions in terms of reduced grey and white matter intensity in the middle and medial temporal regions. As revealed in Figure 6, we also employed a less stringent FDR-correction method in our VBM analysis and show the group differences in grey and white matter intensity using both FDR and FWE thresholds. The use of a cluster extent threshold of *P* < 0.001 FDR-corrected involved larger brain regions as shown in the superimposed maps. Importantly, reduced grey and white matter intensity appeared to be asymmetric (L > R) and the right hippocampus was found to be preserved in both maps for the lvPPA group.

**Figure 6.**
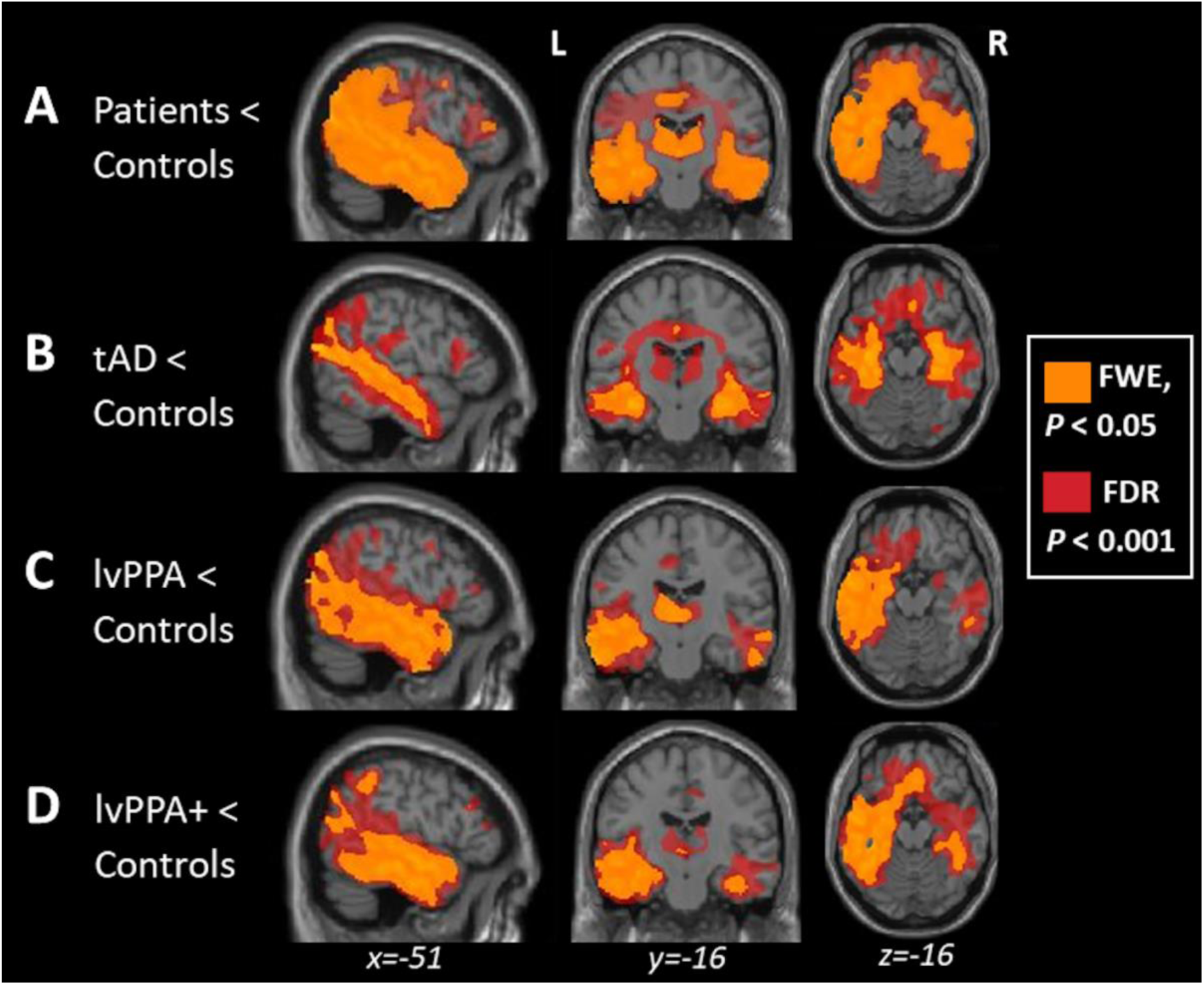
Superimposed voxel-based morphometry results comparing Family-Wise Error- and False Discovery Rate-correction methods. Panels indicate regions of significant grey and white matter intensity reduction in (**A**) all patients (N = 27), (**B**) tAD (N = 9), (**C**) lvPPA (N = 10), and (**D**) lvPPA+ (N = 8) patients compared to controls (N = 12). Voxels in yellow and red indicate regions that emerged as significant at *P* < 0.05 and *P* < 0.001 corrected for Family-Wise Error and False Discovery Rate, respectively, with a cluster threshold of 100 contiguous voxels.

## Discussion

Our results provide strong evidence that individuals with lvPPA exhibit a semantic control deficit across both verbal and non-verbal domains. In contrast to tAD patients who were relatively preserved across semantic tasks, lvPPA patients presented with varied levels of performance across different semantic tests, showing impairments on semantic tests that are more challenging (e.g., alternative object use, synonym judgement task) while being preserved in less-demanding tasks (e.g., word-picture matching). Importantly, relative to controls and tAD patients, lvPPA and lvPPA+ patients were also impaired on non-verbal semantic tests such as the picture version of the Camel and Cactus Test (CCT) and the alternative object use task.

These findings are consistent with prior reports of semantic control deficits in patients with SA or WA patients, further strengthening the proposal that semantic control mechanisms are domain-general and occur across modalities.^5,9,54,65,66^ Like previously reported SA patients, lvPPA and lvPPA+ patients showed positive effects of phonemic cueing and displayed positively correlated performance for semantic control and executive tasks. The only difference between SA and lvPPA patients was that, in the present study, we found strong evidence for an effect of familiarity in the whole group, including tAD and lvPPA patients, even though there was no evidence for an interaction between familiarity and group. We examined the progression of verbal and non-verbal semantic deficits in lvPPA across disease severity by subgrouping the lvPPA patients into two groups, one meeting current consensus criteria (lvPPA N = 10) and the other exhibiting additional cognitive impairments due to disease progression (lvPPA+ N = 8).

Our findings highlight two key issues. Like the post-stroke SA and WA patients, (i) lvPPA patients’ performance declines in line with increasing semantic control demands, and (ii) this pattern of decline is true for both verbal and non-verbal semantic tasks. Results of whole-brain voxel-based morphometry (VBM), comparing each of the patient groups to age-matched controls, were consistent with expectations: the tAD group had significantly reduced grey and white matter in the bilateral hippocampi and lateral temporal regions, whereas the reduced grey and white matter observed in the lvPPA group was largely restricted to the language-dominant left hemisphere, including a large swathe of the temporal lobe. The lvPPA+ group further demonstrated reduced grey and white matter in the left temporal lobe extending subcortically, anteriorly, and posteriorly into the parietal lobule, as well as involvement of right hemisphere temporal regions. In the following sections, we revisit our aims, namely semantic cognition, and imaging patterns in lvPPA relative to tAD patients, interpret our findings, and consider their clinical implications.

### Semantic control in lvPPA

The results of this study indicate that individuals with lvPPA have impaired semantic performance, but not to the degree seen in SD. The lvPPA cohort afforded the opportunity to assess semantic performance across differing levels of severity, comparing mild patients who met strict consensus criteria^11,42^ and those who had progressed with multi-domain impairments over and above logopenia. In addition to the expected poor performance on confrontation naming (one of the two hallmark features of lvPPA), there was strong evidence overall that semantic performance was impaired in lvPPA and lvPPA+ patients relative to controls and patients with tAD, particularly on tests with increased semantic control demands (i.e., Camel and Cactus Test, alternative object use, synonym judgement task). In contrast to lvPPA patients who were at ceiling on the easy CSB word-picture matching task, the lvPPA+ group were impaired. This is in line with previous studies reporting that as lvPPA patients progress, they also present with single word comprehension deficits.^15–20^ In sum, both lvPPA and lvPPA+ patients showed impaired semantic performance. All semantic tests rely on both semantic representations and control, and how much executive-semantic control is required depends on the task (e.g., high for alternative object use task and low for word-picture matching). Whereas the deficits of lvPPA patients were largely restricted to semantic tests with high control demands, lvPPA+ patients exhibited additional emerging deficits on tests with low control demands – a pattern that has been observed previously in SA patients of differing severities.^4,6,67^

The presence of semantic deficits in lvPPA patients raises the question of whether the impaired performance is similar to that of SA and WA cases or of svPPA/SD. The features of semantic control deficits from prior studies of SA and WA cases include inconsistent performance across different types of task, declining performance in line with the control demand levels of the test, no evidence for a significant impact of frequency/familiarity, positive effects of cueing, and positively correlated performance between executively and semantically demanding tasks. In contrast, SD patients have shown consistent item-specific consistency across semantic tasks due to degradation of central semantic representations, strong effects of frequency/familiarity, insensitivity to cueing, and no correlation between executive and semantic task performance. In fact, higher cognition including executive control is reported to be largely spared in svPPA/SD.^68^ The lvPPA patients in our study exhibited varying performance according to semantic control demands across verbal and non-verbal task modalities. Like previously reported SA cases, we found a positive effect of phonemic cueing on naming accuracy in all patient groups. Even in the severely anomic lvPPA+ group, phonemic cueing improved naming abilities, highlighting the idea that when lexical access is impaired, the efficiency of access is boosted by external support. While Cerbone *et al*. similarly found a positive effect of phonemic cueing on BNT performance across AD patients of varying disease severity, the mild AD benefited significantly more than moderately impaired AD patients.^69^ Whether the effect of phonemic cueing may be modulated by disease severity in lvPPA deserves further investigation. As shown in Figure 5, evidence for a correlation between ‘executive’ principal component scores and performance on more demanding semantic control tasks (i.e., Camel and Cactus Test, synonym judgement task) was moderate to very strong in the lvPPA/lvPPA+ combined group, and this correlation was not observed in tAD patients. Taken together, the semantic performance profiles in both lvPPA and lvPPA+ groups mirrored the SA picture in all ways except one. Compared to previously reported svPPA/SD patients who showed robust effects of familiarity and frequency,^4,70^ all groups showed a familiarity/frequency effect which did not interact with group.

Comparisons of verbal and non-verbal modalities in the present study offer valuable insights about the influence of language impairment on semantic performance in lvPPA. If lvPPA is a language-specific clinical entity (i.e., with other cognitive functions intact), then we might expect the semantic impairment in this disorder to be limited to verbal tasks, much like the early ‘semantic access’ patients.^71–74^ Many prior studies have shown that SA and WA patients present with heteromodal semantic deficits in both verbal and non-verbal semantic tasks and, similar to these cases, the lvPPA and lvPPA+ patients in the present study showed impaired performance on non-verbal as well as verbal semantic tests further highlighting the domain-general nature of semantic control mechanisms.^6,9,54^ The alternative object use task is a non-verbal test that has previously been reported to be sensitive in detecting deficits in controlled semantic processing.^5,6,54,60^ Thompson *et al.* found that both SA and WA patients presented with semantic control problems in the non-verbal domain despite having varied lesion profiles with overlap in the posterior temporal region.^9^ The two key takeaways of our results are that (1) patients with lvPPA have core non-verbal deficits alongside their language impairments and (2) the posterior temporal region may be a key semantic control region given the overlap between lvPPA, SA and WA patients.

### Neuroimaging of lvPPA

Our VBM analysis comparing lvPPA patients to controls confirmed the asymmetric pattern of atrophy reported in numerous studies.^18,75–81^ Reduced grey and white matter intensity was largely restricted to the language-dominant left hemisphere within the temporal lobe, both laterally and medially involving the hippocampus and surrounding regions, and parietal regions including the angular gyrus. Similar to the findings of Mesulam *et al*., mediotemporal grey and white matter reductions were confined to the left hemisphere even when using a more lenient threshold as shown in Figure 6.^82^ Only a handful of studies to date have tracked the longitudinal patterns of atrophy in lvPPA.^39,81^ The pattern of atrophy observed in our lvPPA+ group is in line with Rohrer *et al.* who showed that atrophy remains asymmetrical (L > R) over time with increasing involvement of more anterior fronto-temporal areas, as well as right hemisphere regions including the temporoparietal junction, posterior cingulate, and precuneus. Rohrer *et al*. postulated that the emergence of single word comprehension deficits in lvPPA may be related to increasing atrophy in the anterior temporal lobe.^39^ Indeed, patients in the lvPPA+ group showed greater anterior temporal lobe grey and white matter reductions and presented with single word comprehension deficits in the present study.^37,83^

Prior studies have proposed that semantic cognition involves interactions between the anterior temporal lobes (ATLs), serving as a multimodal semantic representation hub, and modality specific spoke regions distributed throughout the cortex.^1,84,85^ The semantic control network is thought to be comprised of the inferior frontal gyrus (IFG) and the lateral posterior temporal cortex, particularly the posterior middle and inferior temporal gyri bounded by the superior temporal sulcus and the fusiform gyrus.^35^ Even though the majority of SA cases have damage to the left IFG, extending to posterior temporal and inferior parietal regions, some SA patients have been reported to have lesions in the temporoparietal cortex with the left prefrontal regions spared much like the lvPPA cases in the present study.^9,86^ WA is also typically associated with lesions in the superior temporal gyrus and the surrounding perisylvian region.^9,87^ As shown in Figure 6, all patient groups had varying degrees of atrophy in the semantic control and representation regions. In the lvPPA group, atrophy encroached most of the left lateral posterior temporal semantic control region as well as a significant portion of the left ATL. Their atrophy profiles overlap with previously reported SA cases in the middle and inferior temporal regions, as well as with the WA cases in the superior temporal and inferior parietal regions. In the more impaired lvPPA+ group, there was atrophy in the left lateral posterior temporal cortex and the left ATL, including the temporal pole, as well as in a significant portion of the right temporal lobe. The ATL involvement in both lvPPA and lvPPA+ patients is noteworthy given that degradation of conceptual representations in svPPA/SD follows atrophy in the ATLs.^2,88–90^ We did not find grey or white matter reductions in the IFG for our lvPPA cohort, which supports previous findings by Thompson *et al*.^86^ that damage to temporoparietal cortex is sufficient to impair semantic control in SA cases. Moreover, while our findings support the proposal by Jackson^35^ that the lateral posterior temporal cortex may be a key semantic control region serving as an intermediary between frontal control and temporal semantic regions, the specificity of anterior versus posterior regions of the superior, middle, and inferior temporal gyri in subserving controlled semantic cognition deserves further investigation.

Although not the focus of the current study, one further question that deserves discussion is the involvement of right mediotemporal regions in lvPPA disease course and the emergence of episodic memory deficits. Mesulam *et al.* proposed that the preservation of episodic memory in lvPPA might be due to the unilaterality of mediotemporal degeneration. Even though our findings are in line with previous studies showing an asymmetrical pattern of atrophy in the lvPPA group, the more progressed lvPPA+ patients in our sample showed right mediotemporal grey and white matter reductions.^37,83^ Moreover, a less stringent threshold of FDR-correction *P* < 0.001 revealed a reduction in a significant portion of the right mediotemporal regions compared to controls. Longitudinal studies of lvPPA with detailed neuropsychological assessments and imaging will be useful in understanding how the spread of atrophy relates to episodic memory and other cognitive functions.

### Limitation

The main limitation to our study is that we were only able to rely on clinical, not pathological, diagnoses. However, clinico-pathological correlations are high in lvPPA. Also, our sample size was relatively small, though we mitigated this by using Bayesian statistics with evidentiary thresholds, linked with detailed neuropsychological assessment and structural MRI.

### Clinical implications

Our study has important clinical implications. There is controversy about whether lvPPA is and remains a language predominant clinical entity over time or progresses to multi-domain dementia.^82,91,92^ The verbal and non-verbal semantic deficits reported here present yet more evidence that it is not just language that is affected in lvPPA. Our findings question the classical view of AD subtypes encapsulated within categorical boundaries (e.g., lvPPA, posterior cortical atrophy, typical-amnestic AD, etc. as specific disorders) and instead support a growing number of recent reports of graded variations within and between the AD phenotypes, which highlight the idea that all of the AD subtypes are positions within a multidimensional space.^30,32,33,92–94^

The positive effect of phonemic cueing on naming ability supports the growing body of literature showing the efficacy for at least some forms of PPA of lexical retrieval treatment utilising phonemic cues. The fact that even lvPPA+ patients benefitted from cueing offers new insights that lexical retrieval treatments^95,96^ may be a viable treatment option for lvPPA patients across varying severity. Anomia is a core feature of PPA variants, as well as tAD to a lesser extent. Early detection of anomia in tAD patients may lead to improved clinical characterisation and appropriate interventions such as speech-language therapy. As such, the selection and choice of assessment should be sensitive and difficult enough to capture the mild naming deficits in tAD.

The present study offers an improved and nuanced clinical characterisation in the lvPPA trajectory of semantic decline: (1) lvPPA patients’ performance declines in line with increasing semantic control demands, and (2) this pattern of decline is true for both verbal and non-verbal semantic tasks. These findings can potentially aid in diagnostic subtyping of PPA subtypes, as well as across disease severity.

## Conclusion

This study establishes the presence of deficits in semantic control in lvPPA, that is, decreasing performance in line with increasing task demands of semantic control, whether the task is verbal or nonverbal. We predicted this outcome given that the atrophy patterns of lvPPA patients overlap with the lesion profiles in semantic aphasia and Wernicke’s aphasia, two patient groups previously demonstrated to have impaired semantic control. Although lvPPA is considered a language disorder, these results indicate that cognitive abilities outside the domain of language are affected. The graded distinctions amongst typical and atypical language phenotypes of AD suggest a dimension of semantic cognition that is not uniquely impaired in semantic dementia but may occur even as a result of Alzheimer’s disease.

## Data availability

The authors confirm that the data supporting the findings of this study are available within the article and its Supplementary material. Participant data and MRI scans may be available, subject to a data sharing agreement required to protect confidentiality and adhere to consent forms.

## Acknowledgements

We sincerely thank our patients and their families for supporting this work.

## Funding

This work and the corresponding author (SKH) were supported and funded by the Bill & Melinda Gates Foundation, Seattle, WA, and Gates Cambridge Trust (Grant Number: OPP1144). This study was supported by the Cambridge Centre for Parkinson-Plus; the Medical Research Council (MC_UU_00030/14; MR/P01271X/1; MR/T033371/1); the Wellcome Trust (220258); the National Institute for Health and Care Research Cambridge Clinical Research Facility and the National Institute for Health and Care Research Cambridge Biomedical Research Centre (BRC-1215-20014; NIHR203312); an intramural award (MC_UU_00005/18) to the Medical Research Council Cognition and Brain Sciences Unit; and Medical Research Council Career Development Award (MR/V031481/1). For the purpose of open access, the author has applied a CC BY public copyright license to any Author Accepted Manuscript version arising from this submission. The views expressed are those of the authors and not necessarily those of the NHS, the NIHR or the Department of Health and Social Care.

## Competing interests

The authors report no competing interests.

## Supplementary material

**Supplementary Table 1.** *Post hoc* tests for Bayesian ANOVAs across semantic tasks.

| Task | Groups |  | Prior Odds | Posterior Odds | BF<sub>10,U</sub> | error % |
| --- | --- | --- | --- | --- | --- | --- |
| Boston naming test | Control | tAD | 0.414 | 10397.104 | 25100.829 | 3.174x10<sup>-7</sup> |
|  |  | lvPPA | 0.414 | 807223.658 | 1.2949x10<sup>+8</sup> | 1.525x10<sup>-9</sup> |
|  |  | lvPPA+ | 0.414 | 9.275x10<sup>+7</sup> | 2.239x10<sup>+8</sup> | 3.339x10<sup>-10</sup> |
|  | tAD | lvPPA | 0.414 | 70.365 | 169.876 | 1.225x10<sup>-5</sup> |
|  |  | lvPPA+ | 0.414 | 11080.967 | 26751.822 | 4.390x10<sup>-8</sup> |
|  | lvPPA | lvPPA+ | 0.414 | 2.151 | 5.194 | 7.215x10<sup>-5</sup> |
| Cambridge Semantic Battery (CSB) naming | Control | tAD | 0.414 | 1.765 | 4.262 | 1.317x10<sup>-4</sup> |
|  |  | lvPPA | 0.414 | 58.001 | 140.026 | 1.426x10<sup>-5</sup> |
|  |  | lvPPA+ | 0.414 | 5620.969 | 13570.220 | 9.795x10<sup>-7</sup> |
|  | tAD | lvPPA | 0.414 | 5.261 | 12.702 | 5.870x10<sup>-5</sup> |
|  |  | lvPPA+ | 0.414 | 252.768 | 610.237 | 7.815x10<sup>-6</sup> |
|  | lvPPA | lvPPA+ | 0.414 | 3.881 | 9.371 | 5.310x10<sup>-5</sup> |
| CSB word-picture naming | Control | tAD | 0.414 | 0.432 | 1.043 | 0.003 |
|  |  | lvPPA | 0.414 | 1.274 | 3.075 | 0.009 |
|  |  | lvPPA+ | 0.414 | 4.640 | 11.203 | 3.706x10<sup>-5</sup> |
|  | tAD | lvPPA | 0.414 | 0.347 | 0.839 | 0.003 |
|  |  | lvPPA+ | 0.414 | 1.205 | 2.908 | 0.007 |
|  | lvPPA | lvPPA+ | 0.414 | 0.432 | 1.043 | 0.003 |
| Camel and Cactus Test | Control | tAD | 0.414 | 4.125 | 9.959 | 1.343x10<sup>-5</sup> |
|  |  | lvPPA | 0.414 | 38.315 | 92.501 | 5.373x10<sup>-6</sup> |
|  |  | lvPPA+ | 0.414 | 217.002 | 523.889 | 4.932x10<sup>-6</sup> |
|  | tAD | lvPPA | 0.414 | 0.323 | 0.779 | 0.002 |
|  |  | lvPPA+ | 0.414 | 2.780 | 6.712 | 3.086x10<sup>-5</sup> |
|  | lvPPA | lvPPA+ | 0.414 | 0.677 | 1.635 | 0.005 |
| Synonym judgement | Control | tAD | 0.414 | 2.003 | 4.835 | 1.003x10<sup>-4</sup> |
|  |  | lvPPA | 0.414 | 44.073 | 106.402 | 1.770x10<sup>-5</sup> |
|  |  | lvPPA+ | 0.414 | 8.381 | 20.235 | 1.938x10<sup>-5</sup> |
|  | tAD | lvPPA | 0.414 | 0.321 | 0.776 | 0.002 |
|  |  | lvPPA+ | 0.414 | 0.994 | 2.399 | 0.005 |
|  | lvPPA | lvPPA+ | 0.414 | 0.527 | 1.273 | 0.003 |
| Alternative object use task | Control | tAD | 0.414 | 0.163 | 0.394 | 0.001 |
|  |  | lvPPA | 0.414 | 23.539 | 56.828 | 4.764x10<sup>-5</sup> |
|  |  | lvPPA+ | 0.414 | 2.514 | 6.070 | 1.907x10<sup>-4</sup> |
|  | tAD | lvPPA | 0.414 | 8.860 | 21.389 | 1.574x10<sup>-5</sup> |
|  |  | lvPPA+ | 0.414 | 1.314 | 3.172 | 0.006 |
|  | lvPPA | lvPPA+ | 0.414 | 0.241 | 0.581 | 0.001 |

The first and second columns indicate the task and the groups being compared. The posterior odds (shown in the fourth column) have been corrected for multiple testing by fixing to 0.5 the prior probability that the null hypothesis holds across all comparisons (Westfall, Johnson, & Utts, 1997). Individual comparisons are based on the default *t*-test with a Cauchy (0, $r=1/\sqrt{2}$ ) prior. As shown in the fifth column, the “U” in the Bayes factor denotes that it is uncorrected. The final column shows the numerical error of the Bayes factor computation.

**Supplementary Table 2.** Voxel-based morphometry results showing group differences in grey and white matter intensity. Voxel-wise differences of grey and white matter intensity between each of the patient versus control groups were assessed using independent *t*-tests, with age and total intracranial volume included as nuisance variables. Clusters were extracted, corrected for family-wise error at *p* < 0.05, with a cluster threshold of 100 contiguous voxels.

| Comparison | Regions | Hemisphere | Number of Voxels | Peak MNI coordinates |  |  | t-value |
| --- | --- | --- | --- | --- | --- | --- | --- |
| All patients < controls | Temporal lobe including lateral and medial regions and the insula | Left | 149799 | -50 | -34 | 0 | 12.04 |
|  | Middle frontal gyrus | Right | 503 | 44 | 12 | 32 | 5.70 |
|  | Parietal lobe including the angular gyrus and inferior parietal regions | Right | 473 | 39 | -60 | 40 | 5.84 |
|  | Inferior parietal lobule | Right | 166 | 51 | -44 | 45 | 5.46 |
|  | Postcentral gyrus | Right | 130 | 39 | -28 | 45 | 5.18 |
|  | Anterior superior temporal gyrus | Right | 116 | 32 | 20 | -28 | 5.81 |
| tAD < controls | Medial and lateral temporal including the hippocampus, fusiform and middle temporal gyri | Left | 23323 | -34 | -16 | -16 | 7.60 |
|  | Medial and lateral temporal including the parahippocampal, fusiform and middle temporal gyri | Right | 11845 | 32 | -24 | -18 | 7.35 |
|  | Superior medial frontal | Left | 204 | -8 | 36 | 40 | 5.90 |
|  | Medial frontal | Right | 103 | 8 | 32 | -16 | 5.36 |
|  | Insula | Right | 131 | 42 | 0 | 6 | 5.66 |
| lvPPA < controls | Temporal lobe including medial and lateral regions, particularly the middle temporal gyrus, extending posteriorly into the parietal and occipital regions | Left | 31376 | -66 | -21 | -8 | 8.83 |
|  | Inferior temporal lobe including the fusiform gyrus | Right | 567 | 57 | -20 | -27 | 6.45 |
|  | Middle temporal gyrus | Right | 282 | 66 | -12 | -10 | 6.10 |
|  | Inferior temporal gyrus | Right | 137 | 58 | -38 | -20 | 5.55 |
|  | Medial frontal gyrus | Left | 112 | -6 | 40 | 33 | 5.75 |
| lvPPA+ < controls | Temporal lobe extending subcortically and medially, including hippocampus, anteriorly, and posteriorly into the parietal and temporo-occipital lobes | Left | 31212 | -42 | -22 | -18 | 8.92 |
|  | Medial temporal, including hippocampus, fusiform, and parahippocampal gyri | Right | 2019 | 39 | -20 | -22 | 6.48 |
|  | Inferior parietal lobule including the supramarginal gyrus | Left | 769 | -50 | -44 | 48 | 6.59 |
|  | Middle frontal gyrus | Left | 257 | -46 | 27 | 26 | 5.56 |
|  | Parietal lobe | Left | 204 | -27 | -57 | 42 | 5.76 |
|  | Middle temporal gyrus | Right | 107 | 56 | -30 | -3 | 5.52 |

## Notes

### Competing Interest Statement

The authors have declared no competing interest.

### Author Declarations

All participants provided written informed consent. Study ethics approval was obtained from the University of Cambridge (REC#01/8/94).

### Summary of Updates

This version has been revised based on reviewer comments during peer review.

